# Chromosome 20p11.2 deletions cause congenital hyperinsulinism via the likely disruption of *FOXA2*

**DOI:** 10.1101/2023.08.16.23294161

**Authors:** Thomas W Laver, Matthew N Wakeling, Richard C Caswell, Benjamin Bunce, Daphne Yau, Jayne AL Houghton, Jasmin J. Hopkins, Michael N Weedon, Vrinda Saraff, Melanie Kershaw, Engela M Honey, Nuala Murphy, Dinesh Giri, Stuart Nath, Ana Tangari Saredo, Indraneel Banerjee, Khalid Hussain, Nick DL Owens, Sarah E Flanagan

**Affiliations:** Department of Clinical and Biomedical Science, University of Exeter Medical School, Exeter, UK; The Genomics Laboratory, Royal Devon University Healthcare NHS Foundation Trust, Exeter, UK; Department of Paediatric Endocrinology, Royal Manchester Children’s Hospital, UK; Department of Paediatric Endocrinology and Diabetes, Birmingham Women’s and Children’s Hospital, Birmingham, UK; Department of Biochemistry, Genetics and Microbiology, University of Pretoria, Pretoria, South Africa; Department of Paediatric Endocrinology, Children’s University Hospital, Dublin, Ireland; Department of Paediatric Endocrinology, Bristol Royal Hospital for Children, Bristol, UK; Royal Cornwall Hospital, Truro, UK; Division of Endocrinology, Sanatorio Güemes, Buenos Aires, Argentina; Department of Paediatrics, Division of Endocrinology, Sidra Medicine, Doha, Qatar

## Abstract

Persistent congenital hyperinsulinism (HI) is a rare genetically heterogeneous condition characterised by dysregulated insulin secretion leading to life-threatening hypoglycaemia. For up to 50% of affected individuals screening of the known HI genes does not identify a disease-causing variant. Large deletions have previously been used to identify novel regulatory regions causing HI. Here, we used genome sequencing to search for novel large (>1Mb) deletions in 180 probands with HI of unknown cause and replicated our findings in a large cohort of 883 genetically unsolved individuals with HI using off-target copy number variant calling from targeted gene panels. We identified overlapping heterozygous deletions in five individuals (range 3-8 Mb) spanning chromosome 20p11.2. The pancreatic beta-cell transcription factor gene, *FOXA2*, a known cause of HI was deleted in two of the five individuals. In the remaining three, we found a minimal deleted region of 2.4 Mb adjacent to *FOXA2* that encompasses multiple non-coding regulatory elements that are in conformational contact with *FOXA2*. Our data suggests that the deletions in these three patients may cause disease through the dysregulation of *FOXA2* expression. These findings provide new insights into the regulation of *FOXA2* in the beta-cell and confirm an aetiological role for chromosome 20p deletions in syndromic HI.

## Introduction

Deletions that affect large regions of genomic DNA have an important role in both rare and common diseases [1, 2]. Monogenic disease can result when a deletion removes the entire, or part of the coding sequencing of a single disease-causing gene. For example, recessively acting partial or whole gene deletions of *ABCC8* or *HADH* cause isolated persistent congenital hyperinsulinism (HI) [3, 4], a genetically heterogenous condition characterised by severe hypoglycaemia as a result of the inappropriate secretion of insulin [5].

Some deletions that affect multiple genes can cause syndromic disease, with the extent of the deletion impacting on the clinical presentation [6]. The phenotype resulting from these large deletions can be readily explained when the deletion disrupts known monogenic disease genes, for example a deletion on chromosome 11p15 causes HI, enteropathy and deafness with loss of the *ABCC8* gene responsible for the HI and loss of the adjacent gene, *USH1C*, causing the enteropathy and deafness [7]. In other large deletion syndromes where HI is a rare feature, for example partial or full monosomy of the X chromosome causing Turner syndrome [8], and the 9p deletion syndrome [9], the precise genetic mechanism leading to the HI has not been fully determined [10].

Large deletions can also cause disease by unmasking a recessive pathogenic variant in a known disease gene, by affecting a differentially methylated imprinted control region (e.g. Beckwith-Wiedemann syndrome [11]) or by disrupting a non-coding regulatory element that is critical for controlling gene expression [12]. The latter is exemplified in a recent study where genome sequencing identified large (∼4.5kb) overlapping *de novo* deletions within an intronic region of the *HK1* gene in two children with HI. These deletions led to the discovery of a ∼42bp region that is critical for controlling HK1 expression within the insulin-producing pancreatic beta-cell [13].

Given the important contribution of deletions to the aetiology of HI and considering that routine genetic testing only identifies a pathogenic variant in 45-79% of cases [14, 15] we searched for large deletions (>1Mb) in an international cohort of children with genetically unsolved HI. We identified overlapping heterozygous deletions on chromosome 20p11.2 in five individuals (range 3-8 Mb); in two this included the HI gene, *FOXA2*. In the remaining three, the deletion encompassed multiple non-coding regulatory elements that are in conformational contact with *FOXA2* suggesting that they may cause disease by disrupting the regulation of *FOXA2* expression within the pancreatic beta-cell.

## Materials and Methods

We studied 1,063 individuals referred for routine genetic testing for HI. Clinical information was provided at referral using a standardised request form. Follow-up data by case note review were requested for individuals where a large deletion was identified. Informed consent was obtained from each of the parents/carers. This study was approved by the North Wales Research Ethics Committee (517/WA/0327).

### Routine screening of genes known to cause HI

DNA was extracted from peripheral blood leukocytes using standard procedures. Disease-causing variants in at least 12 known HI genes (*ABCC8, CACNA1D, GCK, GLUD1, HADH, HNF1A, HNF4A, INSR, KCNJ11, PMM2, SLC16A1* and *TRMT10A*) were excluded by targeted next generation sequencing in all individuals as described previously [16]. This analysis generated an average of three million reads per sample. In all individuals routine screening by read depth analysis using ExomeDepth [17] excluded partial/whole gene deletions of the targeted genes. Deletions on chromosome X, 9p24 and 16p11.2, which are known to cause HI, were excluded using the off-target reads [8, 9, 18, 19].

### Screening for novel disease-causing deletions

We searched for large heterozygous deletions (>1Mb) in the 180 individuals on whom we had whole genome sequencing. Deletions were called by read depth analysis using SavvyCNV [19] (default parameters, bin size 2kbp, samples segregated by sequencing machine and sex). We searched for overlapping deletions present in at least three individuals with HI. Deletions which appeared in 882 in-house controls were assumed to reflect common variation or an artefact of the screening method and were excluded. Sequencing data was used to fine map the deletion breakpoints with the boundaries determined by manual inspection in the Integrative Genomics Viewer (IGV) [20] based on the boundary of the drop in coverage.

When a deletion was identified replication studies to search for chromosome 20p11.2 deletions were performed in the 883 remaining individuals with HI using off-target next generation sequencing data which was analysed using SavvyCNV [19] (transition probability 0.001, bin size 200kbp, samples segregated by targeting panel and sex). Due to the limitations of off-target copy number variant (CNV) calling the CNV boundaries are only accurate to +/-200kb. To ensure that the novel deletions identified in our cohort were rare in the population we screened 6,574 in-house controls for deletions in the same region using the same method. Then screened for deletions in this region in two population control cohorts: UK Biobank (n=488,377) [21, 22] and the gnomAD structural variant (SV) database v2.1 (n=150,119) [23].

Deletions identified from off-target CNV calling were confirmed by an independent cytogenetics analysis (patient 4) or by digital droplet PCR (ddPCR) (patient 5). ddPCR (Bio-Rad QX200 system) involved an EvaGreen dye-binding assay to measure dosage at 11 target sites across a 6.6Mb region (Chr20:17930867-24565591) which extended across the *FOXA2* gene [24, 25]. Targets and primer sequences are provided in Supplementary Table 1. This analysis also allowed for refinement of the 5’ breakpoint in patient 5.

When a deletion was identified, parental samples were tested by ddPCR (n=3 families) or off-target CNV calling from targeted panel data (n=2 families) [19]. Family relationships were confirmed by microsatellite analysis (PowerPlex kit, Promega, Southampton, UK).

### Interrogation of sequencing data and epigenomic data to decipher disease mechanism

Genome sequencing data was analysed to search for a recessive variant unmasked by the deletion in three probands. To do this we called all non-synonymous variants using an approach based on the GATK best practice guidelines. Briefly, reads were aligned to the GRCh37/hg19 human reference genome with BWA mem (v0.7.15) followed by local re-alignment using GATK IndelRealigner (v3.7.0). Variants were called using GATK haplotype caller and annotated using Alamut Batch (Interactive Biosoftware v1.11, Rouen, France). All variants common in gnomAD 2.1.1 (AC>500) were excluded.

In an attempt to pinpoint the genomic region that is causative of the HI we searched for *de novo* variants within the minimal deleted region in 103 genetically unsolved individuals with HI where genome sequencing data was available on the proband and their unaffected parents. Variants were called using the pipeline outlined above then confirmed as *de novo* by DeNovoCNN [26].

To assess the expression of the genes disrupted by the novel deletion we studied publicly available human islet single-cell RNA-seq (scRNA-seq) datasets collected over a time course of pancreatic differentiation projected onto a differentiation pseudotime obtained from [27]. We identified consistent temporal trends using Gaussian Process regression, following the approach that we have previously applied [13]. For scRNA-seq data in human islets [28], accession GSE101207, we used gene counts per cell for the size healthy donors and normalized by depth per cell. All accessions used in this analysis are provided in Supplementary Table 2.

We next interrogated assay for transposase-accessible chromatin sequencing (ATAC-seq) datasets to identify whether the loss of minimal deleted region has the potential to impact the regulation of *FOXA2* expression. Quantification of genomic single-nucleus ATAC-seq (snATAC-seq), bulk ATAC-seq and chromatin immunoprecipitation followed by sequencing (ChIP-seq) data was performed as in Wakeling *et al*. [13]. For human islet snATAC-seq data from Chiou *et al*. [29] (GSE160472), total snATAC-seq beta-cell peaks were obtained by generating a bam file of all reads assigned to beta_1 cluster (obtained https://github.com/kjgaulton/pipelines/tree/master/islet_snATAC_pipeline), and then calling peaks with MACS2 v2.2.7.1.

To determine the number of distinct active regulatory regions within the *FOXA2* control region, we calculated depth normalised transcription factor occupancy/chromatin accessibility in reads per million of single-end fragments extended 120bp and the paired-end fragments of all islet transcription factor binding data and snATAC data for alpha_1 and beta_1 clusters. We then took the union of all intervals for which at least one dataset exceeded 1.5 RPM. Human islet Hi-C data were obtained from experiment accession TSTSR043623 and file accession DFF064KIG (.hic file) and TSTFF938730 (bedpe file) [30], were processed and visualised as in [13]. EndoC-βH1 RNA Pol II ChIA-PET enhancer promoter loops [31] were obtained from GSM3333915.

## Results

We identified large overlapping deletions on chromosome 20p11.2 in three unrelated probands from our cohort of 180 HI probands with genome sequencing data. These were the only large overlapping deletions present in three or more HI probands that were not present in our in-house controls. To replicate this finding in a larger cohort, we used off-target sequencing data generated from routine genetic testing for HI to screen an additional 883 individuals and identified a further two probands with deletions which overlapped this region. Testing of parental samples confirmed that the deletions had arisen *de novo* in four individuals whilst one child (patient 3) had inherited the variant from their unaffected mother, who was also heterozygous. Samples from the maternal grandparents in this family were not available for testing.

All five deletions encompassed a non-imprinted region on chromosome 20p11.2. Patient 3 had a complex variant with a deletion of 2.1Mb (Chr20:19507014-21588883) followed by a 0.9Mb inversion (Chr20:21,588,883-22,510,428) and a further 15Kb deletion (Chr20:22,510,428-22,525,896). The deletions in the five patients ranged from 3-8Mb and had a minimal shared overlap of 2.4Mbp. This was fine mapped to 2,367,250bp using genome sequencing data from three individuals (Chr20:20,158,646-22,525,896) (Figure 1). No large deletions spanning the minimal deleted region were identified in 6,574 internal controls or >600,000 population controls recruited to the UK Biobank and gnomAD SV.

**Figure 1:**
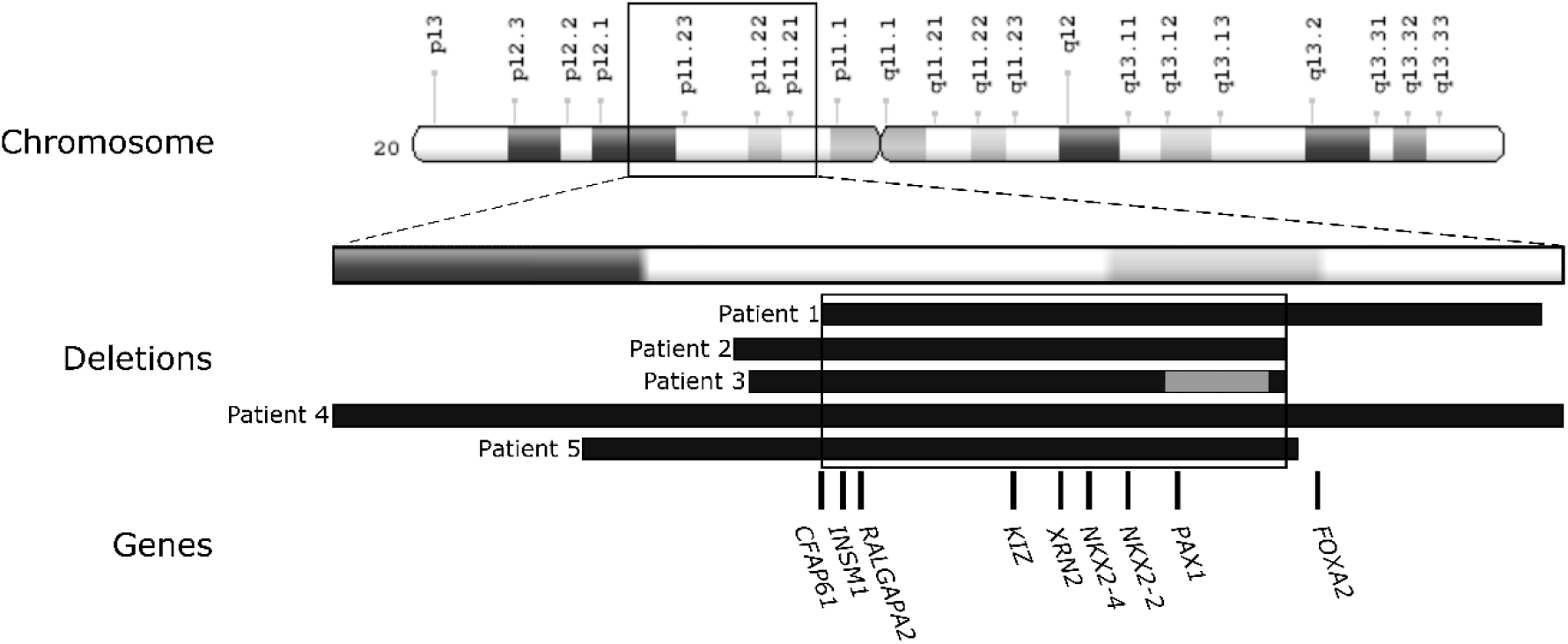
A diagram showing the 20p11.2 deletions in five probands. The chromosomal location of the deletions is at the top. The deletions are depicted by black bars for patients 1-5. A grey bar represents the inversion in the middle of two deletions that was identified in patient 3.

The five probands with a 20p11.2 deletion were diagnosed with HI between the ages of 1 day and 52 weeks. Three individuals were treated with diazoxide while one child showed poor response to treatment necessitating a near-total pancreatectomy. Pancreatic tissue had not been stored following surgery. In one proband the HI had remitted at 6 months, in the remaining three who had not undergone pancreatic resection the HI was ongoing at a median age of 3 years (range 3-12 years). All five individuals had additional features of which two were shared by more than one individual (developmental delay n=4, facial dysmorphism n=3) (Supplementary Table 3).

The ∼2.4Mb shared minimal deleted region contains the entire coding region of 7 genes (*INSM1, RALGAPA2, KIZ, XRN2, NKX2-4, NKX2-2* and *PAX1*). The 5’ boundary dissected *CFAP61*, removing 14 of its 27 exons and the 3’ boundary was located 37kb downstream of the coding region of *FOXA2* (Figure 1). In two individuals the deletion extended over the entire *FOXA2* coding sequence.

To test whether the deletions were unmasking a recessive coding variant we analysed genome sequencing data from three probands, but no rare non-synonymous variants were identified. We also searched for *de novo* variants in genome sequencing data from a further 103 individuals with genetically unsolved HI to see if we could pinpoint the disease-causing gene or regulatory region. No *de novo* non-synonymous variants were detected and no non-coding *de novo* variants within 10kb of each other were found, making them unlikely to be within the same regulatory region.

To investigate the functional impact of the deletions, we assessed the expression of *FOXA2* and the 8 genes within the minimal deleted region whose coding sequence was partially or fully lost. Human islet single-cell RNA-seq data [28] demonstrated that all genes except for *CFAP61, NKX2-4*, and *PAX1* are expressed in islets and pancreatic beta-cells. *FOXA2, NKX2-2, XRN2, INSM1* were most highly expressed (Supplementary Figure 1).

Motivated by the knowledge that heterozygous loss-of-function variants in *FOXA2* cause HI [32-35] and that the deletion in two of the individuals encompassed the entire *FOXA2* gene, we next assessed the gene regulatory potential of the minimal deleted region. Human islet single-nuclei ATAC-seq [29] revealed 149 distinct regions of chromatin accessibility in insulin-secreting beta-cells, including the promoters of the five beta-cell expressed genes (*INSM1, RALGAPA2, KIZ, XRN2* and *NKX2-2*) (Figure 2). We next determined whether any of these regulatory regions had the potential to regulate genes outside the minimal deleted region by examining chromatin conformation by human islet Hi-C data [30]. Amongst multiple three-dimensional contacts, we found a ∼350 kb topologically associated domain spanning *FOXA2* and the minimal deleted region (Figure 2, white arrow). This region has previously been reported to contain islet super enhancers [36-38].

**Figure 2:**
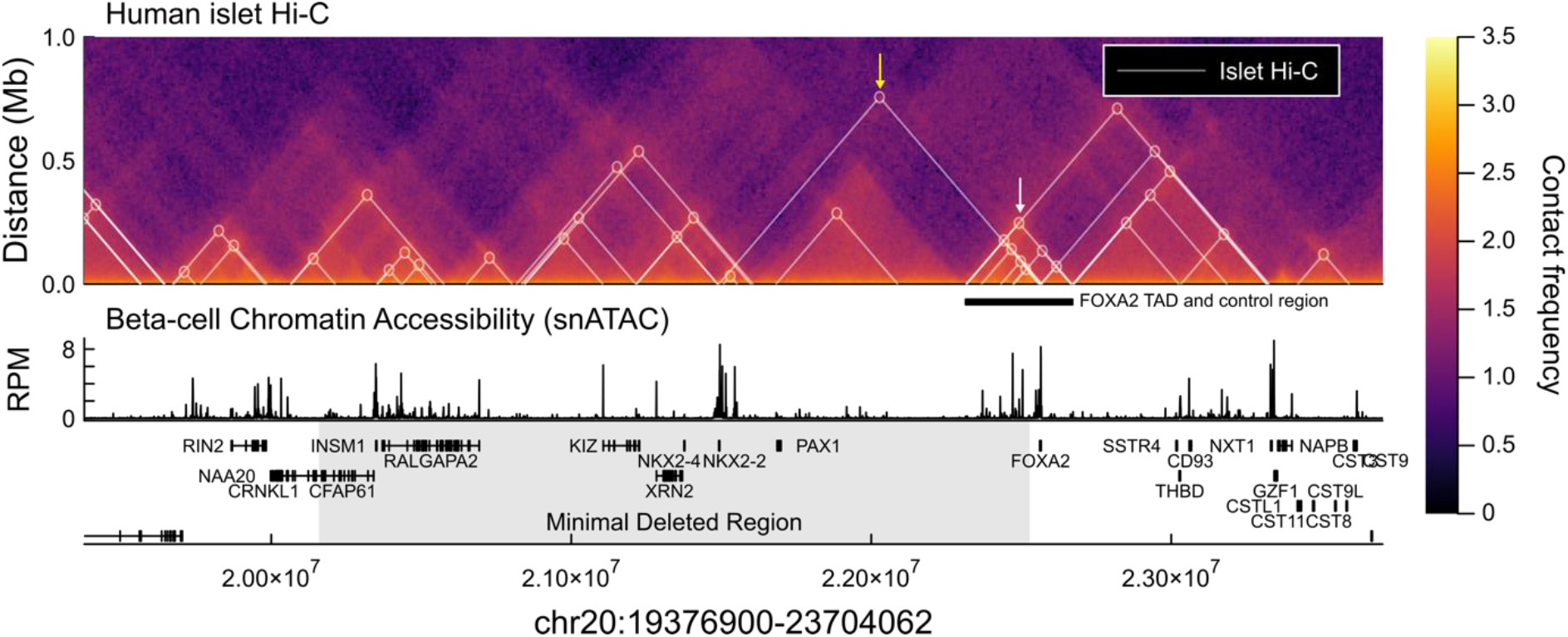
Regulatory landscape of the Chr20 minimal deleted region. Human islet Hi-C contact frequency heatmap [30] (top), and single-nucleus ATAC-seq beta-cell chromatin accessibility [29] (bottom). White triangles and circles mark chromatin loops called in [30]. The white arrow and black bar mark the topologically associated domain (TAD) spanning *FOXA2* and minimal deleted region. The yellow arrow highlights the chromatin loop between *FOXA2* and *NKX2-2*. The grey region marks the minimal deleted region.

Multiple lines of evidence suggest that this region acts as a *FOXA2* control region to regulate expression across differing adult and developing tissues. We identified a 220 kb region (Chr20:22,359,758-22,516,969) encompassing 77 distinct regulatory regions in islets (Supplementary Figure 2). We found that the activity of these regulatory regions varied across pancreatic cell differentiation, between pancreas and liver where *FOXA2* is also highly expressed, and within islet cell types (Supplementary Figure 2). For the latter, the regulatory region most strongly bound by the beta-cell restricted transcription factor PDX1 is accessible in beta-cells and not alpha-cells, suggesting this region confers beta-cell specific control of FOXA2.

Beyond the *FOXA2* control region, human islet chromatin conformation data reveals a CTCF-CTCF loop connecting the promoters of *NKX2-2* and *FOXA2* identifying a co-regulatory mechanism between these two genes that is lost in the minimal deleted region (Figure 2, yellow arrow). It should be noted that *FOXA2* and *NKX2-2* occupy many of the same regulatory loci, with *FOXA2* sharing 45% of its human islet binding sites with *NKX2-*2 [36], and therefore a loss of *NKX2-2* will lead to a disruption of *FOXA2* binding.

Together, our analysis of public epigenomic data indicates that in addition to the loss of coding sequence of 8 genes, the minimal deleted region includes multiple regulatory elements whose loss is predicted to disrupt *FOXA2* expression.

## Discussion

We have identified large overlapping deletions on chromosome 20p11.2 in five individuals with HI. In four the deletions had arisen *de novo* providing strong evidence for pathogenicity. In the fifth proband a complex deletion/inversion variant was identified which had been inherited from their unaffected mother, in keeping with variable penetrance. The absence of rare single-nucleotide variants on the non-deleted allele in three individuals and the lack of an imprinted region suggests that these deletions are causing disease through haploinsufficiency.

Large contiguous gene deletions of varying size on chromosome 20p are well described but relatively rare [39-43]. These deletions have been associated with many developmental and anatomical abnormalities including heterotaxy, biliary atresia, midline brain defects associated with panhypopituitarism, intellectual disability, scoliosis, and seizures. In some cases, specific phenotypic features can be explained when the deletion encompasses a known monogenic disease gene (e.g. *JAG1* deletion causing Alagille syndrome). The five probands in our cohort had extra-pancreatic features which have all been individually described in children with the chromosome 20 deletion syndrome.

All five probands in this study were referred for genetic testing for HI, a condition not commonly associated with chromosome 20 deletions. A literature search identified two cases with HI and growth hormone deficiency caused by heterozygous chromosome 20 deletions. One was a 5.8Mbp deletion on 20p11.22-p11.21 [43] that covers almost the entirety of our minimal deleted region including part of the *FOXA2* control region we identified. The other is a 2.48Mbp deletion on chromosome 20p11.23-p11.21 [40], that overlaps with our minimal deleted region but does not extend over *CFAP61, INSM1* or *RALGAPA2* suggesting that the disruption of these genes is not responsible for the HI.

Considering the overlapping deletion reported by Sugawara *et al* [40] we are left with 5 genes within the minimal deleted region for HI: *KIZ, XRN2, NKX2-4, NKX2-2* and *PAX1*. Convincing data to support a role for each of these genes in the aetiology of HI is lacking. *XRN2, NKX2-4, NKX2-2* and *PAX1* are tolerant to truncating variants based on their gnomAD pLI score (threshold pLI >= 0.9) (data unavailable for *KIZ*) [44, 45] and scRNA-seq data shows that *NKX2-4*, and *PAX1* are not expressed in islets and insulin-producing pancreatic beta-cells. Only *NKX2-2* is known to have a role within the pancreas where it encodes a transcription factor involved in pancreatic cell differentiation, maintenance of beta-cell function and formation of islet structure [21]. Importantly however, bi-allelic null variants in *NKX2-2* cause neonatal diabetes and the parents, who are heterozygous carriers, do not have hypoglycaemia [25]. Recent papers describing *NKX2-2* expression have also shown increased rather than decreased expression in islets from patients with HI [26, 27].

*FOXA2* is in contrast a strong candidate gene for the HI despite the coding sequence of the gene only being deleted in 2/5 individuals in our study and 1/2 individuals previously reported to have HI caused by deletions on chromosome 20p. This gene encodes a transcription factor that has an essential role in pancreatic development [46-49]. In mature pancreatic islet cells, FOXA2 has been shown to regulate the expression of genes that encode key components of the insulin secretion pathway [50, 51]. Most convincingly, heterozygous loss-of-function coding variants in *FOXA2* have been reported in four individuals with HI [32-35] and three individuals with transient hypoglycaemia [34].

In keeping with a role for *FOXA2* in the aetiology of HI in the three patients in our study who have the coding region of the gene intact, our analysis of functional genomic data highlighted a *FOXA2* control region that was deleted in all individuals. This control region has strong three-dimensional contact with the *FOXA2* promoter and comprises at least 77 distinct active regulatory regions within human islets. We find that these regulatory regions exhibit cell-specific regulatory activity, varying in activity across differentiation and adult tissues where *FOXA2* is expressed; it therefore likely plays a role in fine-tuning *FOXA2* expression. Part of this *FOXA2* control region was also deleted in the patient with HI studied by Wee *et al*. [43]. All seven of the patients identified in this study or from the literature who have HI and a deletion on chromosome 20p have either the *FOXA2* coding sequence or at least part of this *FOXA2* control region deleted.

We acknowledge that our patients have deletions affecting significant components of the islet transcription factor network and the phenotypes and impact on gene expression could be multifaceted. Given that *FOXA2* and *NKX2-2* bind many of the same regulatory loci, with *FOXA2* sharing 45% of its human islet binding sites with *NKX2-*2 [27], it also remains possible that concomitant loss of *NKX2-2* is required for HI to develop in these individuals. Furthermore, no *de novo* coding variants or clusters of non-coding variants within the minimal deleted region were identified in a large unrelated cohort of individuals with genetically unsolved HI. This suggests that single nucleotide variants within this region are an extremely rare cause of HI or that large structural variants, that disrupt multiple genes/regulatory regions, are required to cause disease. Identifying further deletions or disease-causing single nucleotide variants that refine the critical region will be important to gain knowledge of the precise molecular mechanism(s) of HI.

In conclusion, we have identified a 2.4Mb deletion on chromosome 20p11.2 as a cause of syndromic HI and we recommend that this chromosome region is included by genomic laboratories in their screening panels for this condition, especially when syndromic disease is suspected. Our findings suggest that these deletions cause HI through the disruption of *FOXA2*, either by removing its entire coding region or by disrupting non-coding regulatory elements that are critical for controlling *FOXA2* expression within the pancreatic beta-cell. These findings further highlight the critical role of studying large structural variants to gain insights into non-coding gene regulation and to aid discovery of novel causes of Mendelian disease.

## Data Availability

All non-clinical data analyzed during this study are included in this article (and its Supplementary Information). Clinical and genotype data can be used to identify individuals and are therefore available only through collaboration to experienced teams working on approved studies examining the mechanisms, cause, diagnosis and treatment of diabetes and other beta cell disorders. Requests for collaboration will be considered by a steering committee following an application to the Genetic Beta Cell Research Bank (https://www.diabetesgenes.org/current-research/genetic-beta-cell-research-bank/). Contact by email should be directed to S. Flanagan. All requests for access to data will be responded to within 14 d. Accession codes and DOI numbers for all ChIP-seq, ATAC-seq, RNA-seq and scRNA-seq datasets are provided in Supplementary Table 2. We used the Genome Reference Consortium Human Build 37 (GRCh37) to annotate genetic data (accession number GCF_000001405.13). Details of this assembly are provided at https://www.ncbi.nlm.nih.gov/assembly/GCF_000001405.13/.

## Acknowledgments

MaNW is a recipient of an Independent Fellowship and TWL and NO a Lectureship from the Exeter Diabetes Centre of Excellence funded by Research England’s Expanding Excellence in England fund. SEF is a Wellcome Trust Senior Research Fellow (223187/Z/21/Z). This study was supported by the National Institute for Health and Care Research Exeter Biomedical Research Centre. The views expressed are those of the author(s) and not necessarily those of the NIHR or the Department of Health and Social Care. This research has been conducted using the UK Biobank Resource. This work was carried out under UK Biobank project number 9055 and 9072. For the purpose of open access, the author has applied a CC BY public copyright license to any Author accepted Manuscript version arising from this submission. We are grateful to Prof. Evgenia Globa for reviewing the presentation of the clinical data reported in this manuscript and to Prof. Amanda Ackermann for providing a clinical update on a patient. The authors have no conflicts of interest to declare.

