## Supplementary Information for "Chromosome 20p11.2 deletions cause congenital hyperinsulinism via the likely disruption of *FOXA2*"

**Supplementary Table 1:** Genomic targets and primers for the droplet digital PCR testing of the 20p.11.2 deletions.

| **GRCh37 genomic coordinates** | **Forward** | **Reverse** |
| --- | --- | --- |
| chr20:17930867-17930972 | TTGTGTGAAAGCTGACAAAATG | CAAGTCTTTTTATCCAAACACAGC |
| chr20:18794590-18794685 | GCTTCCCAGTGTTTCAGGAC | TGGTGATGTCCAGCTGAAAG |
| chr20:19956252-19956341 | ACTTCGGGTGCTTAGTGCAG | GGATGGTCTGCAGCATGTC |
| chr20:21687223-21687326 | GCTGCGTGAGCAAGATCC | CTTGACCACGTTGGGAGTG |
| chr20:22262921-22263030 | AAGGACATGAATGACAAGATC | GAGCTTGATGGAACAATCACC |
| chr20:22381210-22381300 | TTTGCTCCTTTTGACTGCTG | ACCCATGTGTTCATCCACTG |
| chr20:22441027-22441122 | TGAGTGGCAAACACCTGAAC | CAAGGCCTCTTGAGGTATGC |
| chr20:22550245-22550351 | GCCTGGAAATTTGTCTGAGC | TTGCAATGTCTGTGCAGGTC |
| chr20:22564830-22564927 | TTTAAACTGCCATGCACTCG | CTCGGGCTCTGCATAGTAGC |
| chr20:23370596-23370687 | GCTTTGGTTCTTGATTTCAGC | TCAATGGCTTTCTGGTACTGC |
| chr20:24565487-24565591 | TGCAGAGCGACTACTCAAGC | AGAAGCAGCAGAGCATGGAG |

**Supplemental Table 2:** Table describing all public genomic datasets used in this study.
ES – embryonic stem, DE – definitive endoderm, GT – gut tube, PP – pancreatic progenitor.

| Accession | DOI | Description | Cell | Target |
|---|---|---|---|---|
| GSE149148 | doi.org/10.7554/eLife.59067 | ATAC-seq, ChIP-seq for TFs over pancreatic differentiation | ES, DE, GT, PP1, PP2 | FOXA2, ATAC |
| E-MTAB-1919 | doi.org/10.1038/ng.2870 | ChIP-seq for TFs in islets | Islets | CTCF, FOXA2, H2AZ, H3K27ac, MAFB, NKX2_2, NKX6_1, PDX1 |
| E-MTAB-1990, E-MTAB-3061 | doi.org/10.1038/ncb3160 | ChIP-seq for TFs liver buds | LiverBud, | FOXA2 |
| GSE148368 | doi.org/10.1038/s41467-021-26950-0 | ChIP-seq for TFs in pancreatic and liver differentiation | GT, HP | FOXA2 |
| GSE160472 | doi.org/10.1038/s41588-021-00823-0 | Single nuclei ATAC-seq in islets | Islets | ATAC |
| GSE101207 | doi.org/10.1016/j.celrep.2019.02.043 | Single cell RNA-seq in islets | Islets | |
| GSE143783 | doi.org/10.1038/s42255-020-00314-2 | Single cell RNA-seq of beta-like cell differentiation, abundances projected on pseudotime available in ref's Supplementary Table 4 | ES cell to beta-like cell differentiation | |

**Supplementary Table 3:** Summary of clinical features and genetic findings in individuals with 20p11.2 deletions. Genetic coordinates relate to GRCh37. * denotes the total genomic region disrupted in this patient which includes an inverted region (details provided in the main text).

| | Patient 1 | Patient 2 | Patient 3 | Patient 4 | Patient 5 |
|---|---|---|---|---|---|
| **Coordinates of 20p.11.2 deletions** | Chr20:20158646-24080787 | Chr20:19434987-22528253 | Chr20:19507014-22525896* | Chr20:16400000-24400000 | Ch20:18200000-22600000 |
| **Age at Presentation of HI** | <1 week | <6months | <1 year | <1 week | <1 week |
| **Glucose (mmol/L) in the presence of detectable insulin** | ≤3 | ≤3 | ≤3 | ≤3 | ≤3 |
| **Syndromic Features** | Yes | Yes | Yes | Yes | Yes |
| **Additional features shared by more than one individual** | Developmental delay | Developmental delay<br><br>Facial dysmorphism | Developmental delay<br><br>Facial dysmorphism | None | Developmental delay<br><br>Facial dysmorphism |

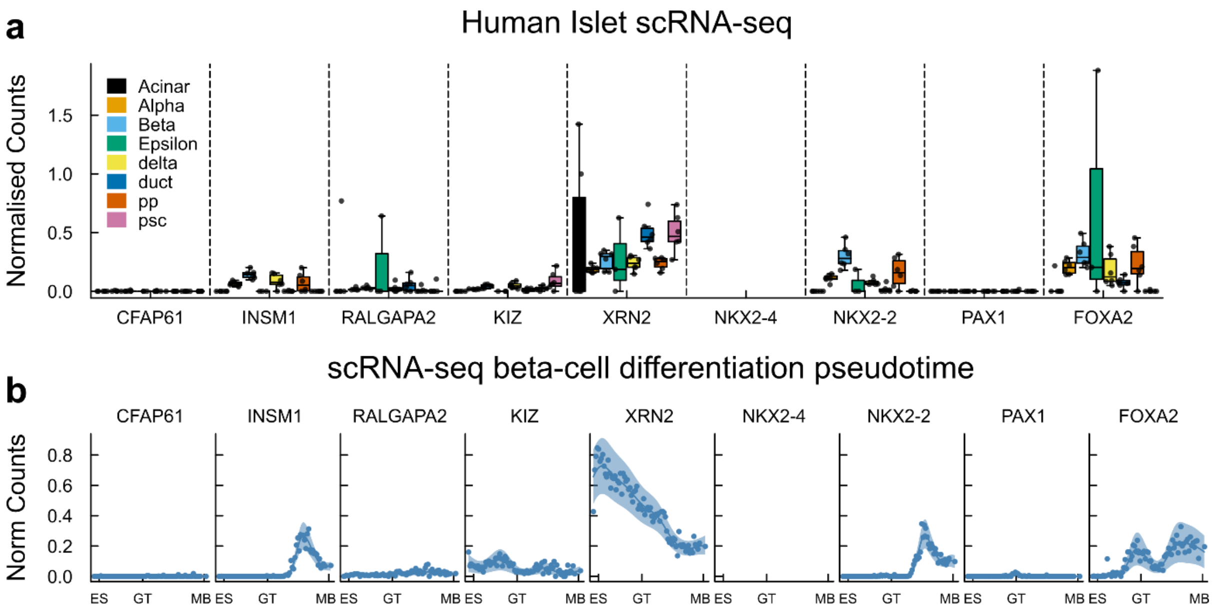


**Supplementary Figure 1:** Expression of genes partially or fully deleted by minimal deleted region.

**a.** Human islet single-cell RNA-seq (scRNA-seq) (GSE101207 [1]) expression data of the 8 genes whose sequence is disrupted and FOXA2. Data points give normalised counts of 6 independent donors. Boxplot central lines give the median, boxes span interquartile range, and whiskers extend to the furthest point within 1.5x IQR of the box. Order of cell types (left to right) matches order of legend (top to bottom).

**b.** scRNA-seq expression over pancreatic cell differentiation from embryonic stem (ES) cells to maturing beta-cells (MB), mapped onto a beta-cell differentiation pseudotime [2] that includes gut tube stage (GT). scRNA-seq data-points on pseudotime shown with Gaussian process regression, line and shaded region mark posterior Gaussian process median and 95% confidence intervals.

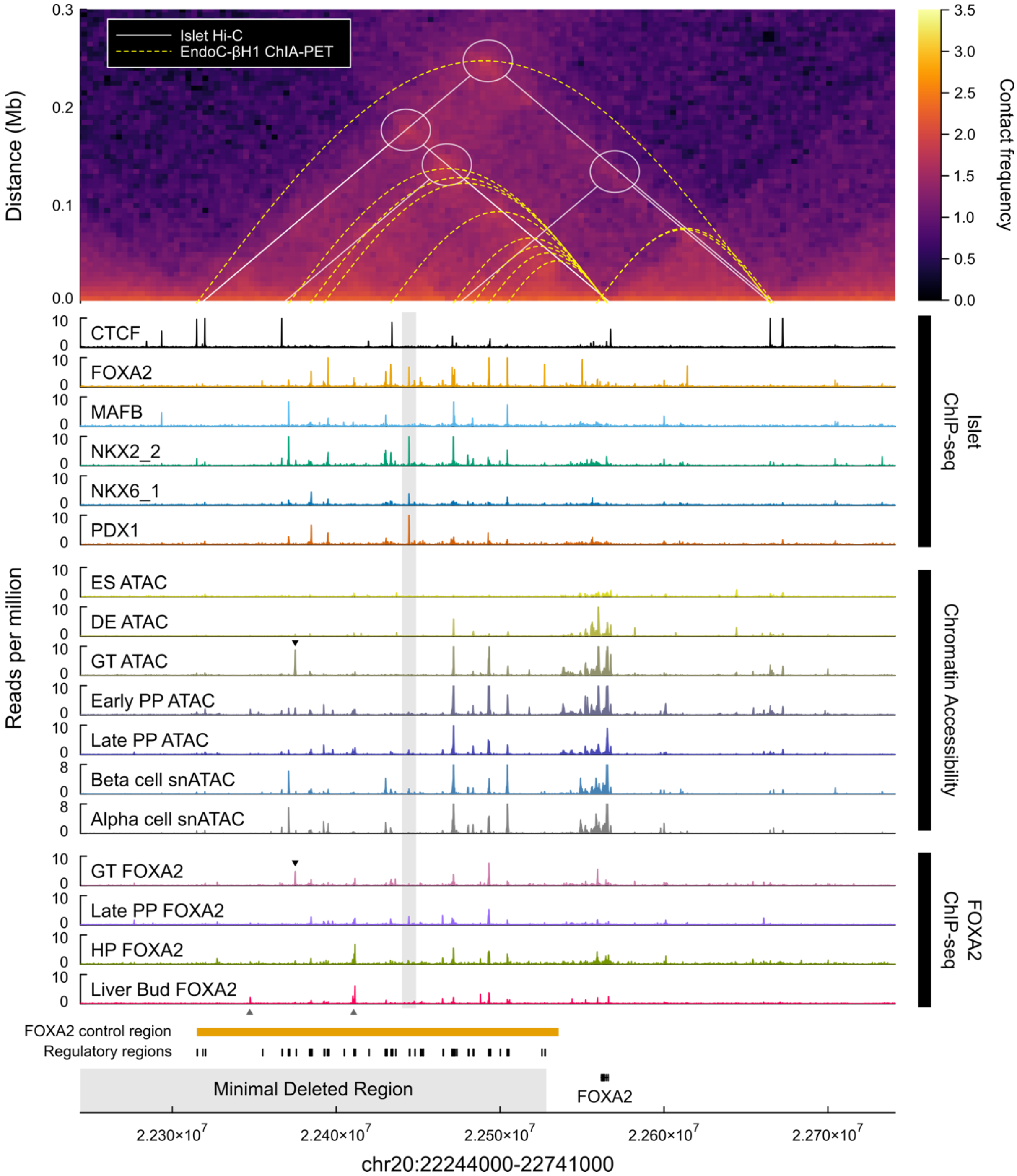


**Supplementary Figure 2:** Regulatory activity of FOXA2 control region.

Human islet Hi-C contact frequencies, transcription factor binding in multiple cell types and chromatin accessibility show regulatory activity of FOXA2 control region (orange bar).

Top: Heatmap gives contact frequencies of human islet Hi-C (5 kb bins) [3], white lines and circles describe CTCF-CTCF chromatin loops called in same study. Yellow dotted lines mark EndoC-βH1 RNA Pol II ChIA-PET enhancer-promoter loops (GSM3333915 [4]), highlighting contact between individual transcription factor binding sites and *FOXA2* promoter.

Bottom: islet chromatin immunoprecipitation followed by sequencing (ChIP-seq) data [5]; chromatin accessibility data over pancreatic cell differentiation [6], ES – embryonic stem, DE – definitive endoderm, GT – gut tube, PP – pancreatic progenitor; single nuclei assay for transposase-accessible chromatin sequencing (snATAC) of human islets showing alpha and beta cell clusters [7], (beta_1 and alpha_1 clusters from original study shown); FOXA2 ChIP-seq from *in vitro* differentiation gut tube (GT), pancreatic progenitors (PP), and hepatic progenitors (HP) [8], and *in vivo* in liver bud [9].

Individual regulatory regions shown strong contact with FOXA2 promoter by Hi-C and RNA Pol II ChIA-PET, activity of regulatory regions varies by factor and cell type. Beta-cell specific regulatory region, marked by grey vertical bar. FOXA2 ChIP-seq shows varied binding of FOXA2 at its own control region across differentiation and between pancreas and liver. Black down triangle (▼) marks gut-tube specific regulatory region by chromatin accessibility and FOXA2 binding. Grey up triangle (▲) marks liver dominant binding of FOXA2.
